# BEYOND GLYCAEMIC CONTROL: SOCIAL DETERMINANTS, PSYCHOLOGICAL BURDEN, AND EARLY COMPLICATIONS MARKERS AMONG YOUNG ADULTS WITH TYPE 1 DIABETES IN SELECT CLINICS IN KENYA

**DOI:** 10.64898/2026.11.11.26367836

**Authors:** Phoebe Wamalwa, Cyrus Mugo, Christine Ngaruiya

**Affiliations:** Global Health Emerging Scholar, Yale University; Pediatric Endocrinologist, Kajiado County Referral Hospital; Visiting Scientist, Kenyatta National Hospital; Senior Research Scientist, Kenyatta National Hospital; Assistant professor, University of Washington; Adjunct Faculty, University of Nairobi; Associate Professor, Director, Stanford EM International Global and Population Health Section (SEMI), Department of Emergency Medicine, Stanford School of Medicine

**Keywords:** Type 1 diabetes, Social determinants, glycemic control, complications, psychological health

## Abstract

**Background:** Young adults with type 1 diabetes (T1D) in low- and middle-income countries experience persistently poor glycaemic control despite improved insulin access. Evidence on the effects of social determinants of health (SDOH) on glycaemic control, complications, and psychological well-being remains limited, particularly in sub-Saharan Africa. We examined relationships between SDOH, psychological health, glycaemic control, and early complication markers among young adults with T1D in Kenya.

**Methods:** We conducted a multicentre cross-sectional study among 164 adults aged 18–34 years attending four diabetes clinics in Kenya. SDOH were assessed using a prespecified, context-informed questionnaire covering six domains. Psychological well-being was assessed using PAID, PHQ-9, and GAD-7. Clinical outcomes included HbA1c, a single urine albumin-to-creatinine ratio (UACR), and lipid profile. Associations between SDOH domains and clinical and psychological outcomes were evaluated using multivariable ordinal and binary logistic regression models adjusted for age, sex, and diabetes duration.

**Findings:** Mean age was 24.96±4.35 years and diabetes duration 7.26±5.70 years. Mean HbA1c was 11.5±3.44%, with only 4.9% achieving <7%. LDL cholesterol was above target in 56.1%, 64.0% had an elevated LDL/HDL ratio, and 28.0% had elevated UACR. Diabetes distress and clinically significant depressive symptoms affected 30.5% and 27.5%, respectively. Moderate-to-high SDOH burden was greatest for economic stability (76.8%), social/community context (52.4%), and food environment (43.9%). All SDOH domains showed positive but non-significant associations with HbA1c. Diabetes distress was associated with economic instability (aOR 1.42, 95% CI 1.17–1.72), healthcare system burden (1.45, 1.12–1.87), neighbourhood/physical environment burden (1.39, 1.07–1.80), and food environment burden (1.30, 1.03–1.64). Anxiety was associated with social/community (1.70, 1.14–2.53) and healthcare system burden (1.68, 1.19–2.36). Economic instability was associated with elevated UACR (1.40, 1.05–1.85).

**Discussion:** Young adults with T1D in Kenya experience substantial social and psychological burden alongside poor glycaemic control and early complication markers. Social disadvantage appears to influence kidney disease complications, and psychological well-being regardless of glycaemic control in our sample, underscoring the need to move beyond glucose-centered care. Poor glycaemic control was pervasive, which likely obviated any detectable differences by SDOH in this population. Ultimately, Integrated care addressing SDOH and mental health may support more sustainable long-term outcomes.

## Background

Diabetes mellitus encompasses a collection of chronic metabolic disorders primarily characterized by elevated blood glucose due to deficiencies in insulin production, action, or both [1]. Type 1 Diabetes (T1D) arises when the body’s immune defense mistakenly attacks the pancreas’ beta cells responsible for producing insulin, leading to its complete deficiency. This form comprises around 5–10% of all diabetes cases globally and remains the most frequent form in children and adolescents, although it can emerge at any stage in life [1]. The global incidence of T1D, particularly in sub-Saharan Africa, has shown a notable rise in recent decades [2].

Young adults living with type 1 diabetes (T1D) in low- and middle-income countries (LMICs) experience disproportionately poor health outcomes despite advances in provision of the recommended basal bolus regimen (multiple daily injections) of insulin. [3,4]. The American Diabetes Association (ADA) recommends individualized glycaemic targets, with an HbA1c of <7% (53 mmol/mol) appropriate for most nonpregnant adults [5]. Social determinants of health (SDOH), including economic stability, education and health literacy, social support, neighborhood environment, food security, and healthcare access, account for a substantial proportion of health disparities in diabetes [6]. These determinants influence self-management behaviors, healthcare utilization, treatment adherence, and quality of life [7,8]. Young adults in Kenya face additional vulnerabilities, including high unemployment, financial dependence [9], and transition out of subsidized free insulin pediatric diabetes programs, placing them at increased risk of poor glycemic control, psychological distress, and early diabetes complications.

Psychological disorders such as diabetes distress, depression, and anxiety are common among adolescents and young adults with T1D and may arise from the substantial daily burden of self-management, fear of hypoglycaemia and long-term complications, and the expectations associated with maintaining glycaemic control [10]. These disorders have each been reported in approximately one-third of young people with T1D in some studies and may adversely affect insulin use, glucose monitoring, dietary behaviours, and engagement with diabetes care [11–13]. Persistent psychological distress is consequently associated with poorer glycaemic control, recurrent diabetic ketoacidosis (DKA), and, in severe cases, increased mortality risk, including through intentional insulin misuse or overdose [10,13].

Alongside psychological morbidity, early microvascular complications may begin to emerge during this period [4]. Diabetic kidney disease is among the most serious complications of T1D and develops through the combined effects of chronic hyperglycaemia, haemodynamic alterations, inflammation, and other cardiovascular and metabolic risk factors [14]. Although clinically established nephropathy generally becomes more prevalent with longer diabetes duration, early abnormalities in urinary albumin excretion may occur in adolescents and young adults and can precede progressive renal impairment [14]. Poor glycaemic control, hypertension, dyslipidaemia, smoking, and longer diabetes duration further increase renal and cardiovascular risk. Despite evidence that intensive diabetes management and multifactorial risk-factor control can delay progression [4], early kidney and cardiovascular risk markers may remain under-recognised in routine care, particularly in resource-limited settings.

Despite the growing burden of T1D among young people in sub-Saharan Africa, evidence on the combined influence of social disadvantage, psychological morbidity, and early complication risk in young adults remains limited. Most available studies have examined glycaemic control, mental health, or chronic complications separately, with little attention to how SDOH interact with these outcomes during the vulnerable transition to adulthood. Data from Kenya are particularly scarce on the relationship between SDOH and early renal and cardiovascular risk markers such as albuminuria and dyslipidaemia. This limits the development of context-specific, integrated models of care that address not only insulin access and glycaemic management, but also the social and psychological factors that may shape long-term outcomes.

To address this evidence gap, we evaluated the relationship between six domains of SDOH, psychological well-being, glycemic control, and early renal and cardiovascular complications, including albuminuria and dyslipidemia, among young adults with T1D receiving care in four Kenyan diabetes clinics.

## Methods

This was a cross-sectional study assessing the relationship between SDOH, psychological well-being, and glycemic outcomes among individuals with type 1 diabetes in Kenya. The study was conducted on young adults aged 18 to 34 years living with type 1 diabetes attending diabetes clinics at four geographically and health-system diverse hospitals representing urban, peri-urban, rural, and semi-arid settings. Different levels of care were also included: a level 6 tertiary referral and teaching hospital in urban Nairobi, the capital city, two level 5 peri-urban facilities which constitute county referral hospitals and a level 4 facility which constitutes a sub-county hospital. These facilities were KNH, Machakos and Kajiado Hospitals, located approximately 65 km and 75 km from Nairobi, and Westland’s hospital respectively. The latter hospitals were included to provide representation of diabetes care at a lower-level facility as shown in figure 1.

**Figure 1:**
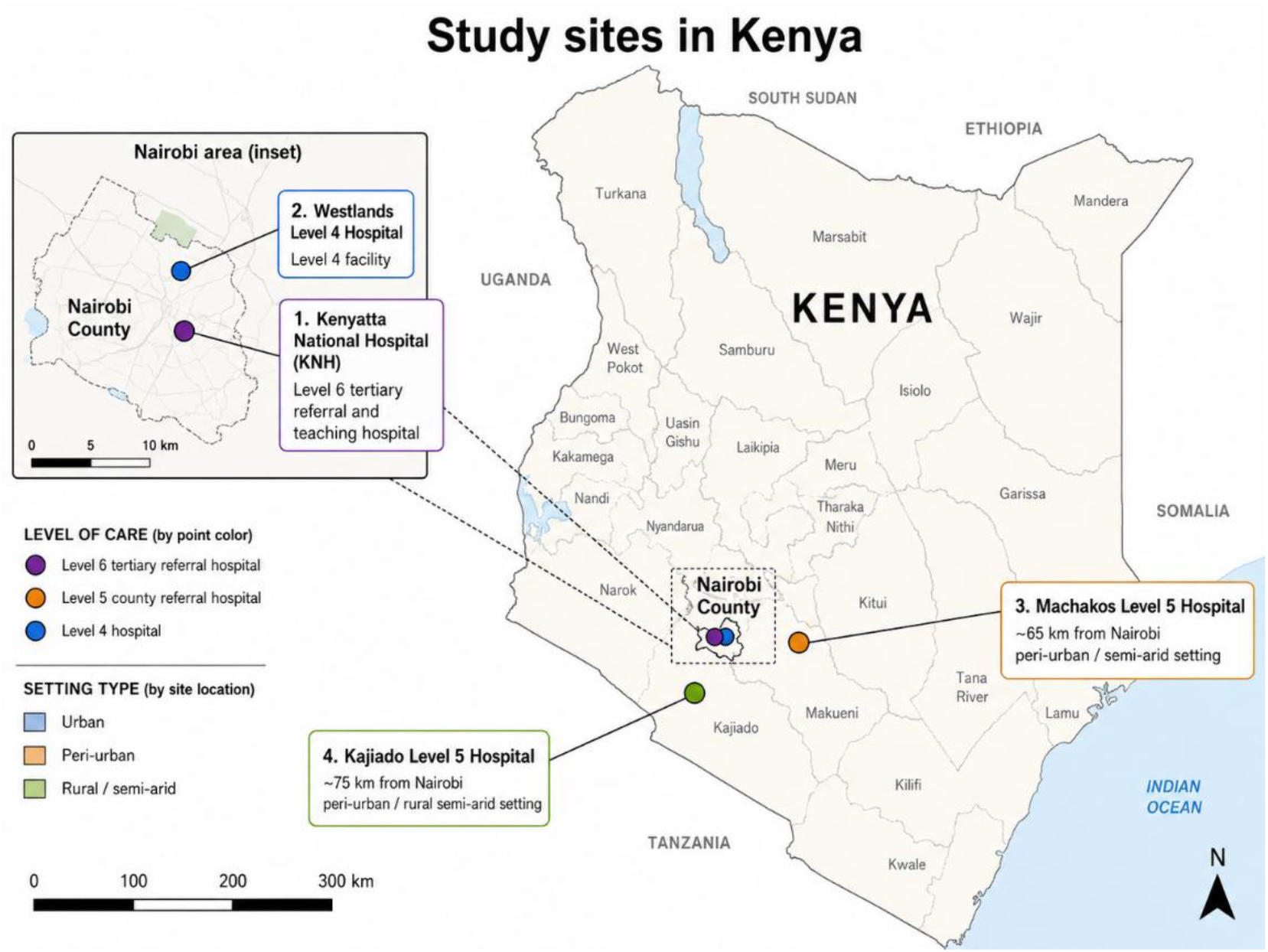
Study site distribution in Kenya. The study included health facilities across different levels of care: Kenyatta National Hospital (KNH), a Level 6 tertiary referral and teaching hospital in urban Nairobi; Machakos and Kajiado County Referral Hospitals, both Level 5 facilities located approximately 65 km and 75 km from Nairobi, respectively; and Westlands Hospital, a Level 4 sub-county facility in Nairobi.

Participants were eligible if they were aged 18–34 years, had a clinician-documented diagnosis of T1D in the medical record, were receiving ongoing insulin therapy, and had a clinical presentation consistent with insulin deficiency [15], including acute onset of diabetes and/or classical hyperglycaemic symptoms at diagnosis. Available medical records were reviewed to support the diagnosis. Conventionally, islet autoantibody and C-peptide testing may be obtained for routine diagnosis in clinical or research studies; however, given these labs are generally inaccessible in our context, a diabetes diagnosis was based on clinical assessment and review of medical records as noted.

The sample size was calculated for multiple regression with six predictors, assuming α=0.05, Cohen’s *f*^2^=0.15, and 95% power, yielding a minimum sample of 150 participants. Although the recruitment target was 188 after allowing for 20% non-response, 164 participants were enrolled, exceeding the minimum required sample. Site-specific recruitment targets were allocated according to the relative clinic caseloads: (Kenyatta National Hospital, n=40; Westland’s Level 4 Hospital, n=23; Machakos level 5 hospital, n=68; Kajiado level 5 hospital, n=33). Eligible participants registered at each clinic were identified through a multimodal recruitment strategy using routine clinic attendance, telephone invitations from hospital records, announcements on diabetes support platforms, and peer referral. Recruitment was conducted by trained research assistants, from September to December 2025, using a consecutive sampling approach, enrolling all eligible participants present during the study recruitment period or identified in records. This approach continued until the target sample for each site was achieved.

Data on SDOH and psychological well-being were collected using a structured questionnaire, while clinical and laboratory assessments captured metabolic and early complication markers. Questionnaire responses were cross-checked against available clinical records.

### Development and contextualization of the SDOH instrument

The SDOH questionnaire was developed before data collection using the Kaiser Family Foundation framework [16], and local contextual evidence [17]. The questionnaire included 18 indicators grouped into six domains, with the domain structure informed by the Kaiser Family Foundation (KFF) social determinants of health framework. Six experts reviewed the items for relevance, clarity, and cultural appropriateness. The revised questionnaire was pretested among 10 young adults with T1D at Kajiado County Referral Hospital who were excluded from the main study. Variations in responses and qualitative feedback from participants were reviewed to identify unclear, ambiguous, or contextually inappropriate items and were used to refine the wording, clarity, and overall usability of the questionnaire before data collection. The tool was administered in English or Kiswahili. The Kiswahili version was translated by a nurse educator, independently back-translated by a bilingual non-medical professional, and discrepancies were resolved by the study team.

### Social determinants of health variables

Social determinants of health (SDOH) variables were prespecified before analysis and grouped into six domains according to the Kaiser Family Foundation framework: economic stability; education and health literacy; social and community context; neighbourhood and physical environment; healthcare access and system experience; and food environment and household food security. Each domain comprised three indicators selected to reflect locally relevant social conditions that could influence T1D self-management as shown in table 1. The Diabetes Index for Social Determinants of Health (DISDOH) [18] was used to support the framing and interpretation of the findings. The full questionnaire and item definitions are provided in the Supplementary Appendix.

**Table 1:** SDOH domains and indicators.

| **Six study SDOH domains (KFF) [16]** | **Constituent indicators** | **Illustrative sample question\*** | **Five domain conceptual DISDOH [18] alignment** |
|---|---|---|---|
| **Economic stability** | Employment/schooling status; income adequacy; household dependency | "What is your current employment or schooling status?" | Socioeconomic status |
| **Education & health literacy** | Educational attainment; diabetes literacy; understanding diabetes-related health instructions | "Do you require assistance to understand diabetes-related health instructions?" | Socioeconomic status; health-literacy |
| **Social & community context** | Social support/support-group participation; frequency of social interaction; companionship/emotional support | "Do you have someone you can talk to or rely on when you need diabetes related support?" | Social context |
| **Neighbourhood & physical environment** | Housing stability/quality and refrigeration; transport barriers; distance to diabetes clinic | "How long does it usually take you to travel to your diabetes clinic?" | Neighbourhood & physical environment |
| **Healthcare access & system experience** | Access to trained/multidisciplinary providers; insurance; appointment adherence/clinic experience | "Do you currently have health insurance coverage?" | Health context |
| **Food environment & household food security** | Food security/food–insulin trade-offs; access to fruits and vegetables; access to snacks at work/school | "Have you ever skipped insulin because you did not have enough food?" | Food environment |

\* *Individual SDOH variables were grouped a priori into six conceptual domains: Economic Stability; Education and Health Literacy; Social and Community Context; Neighbourhood and Physical Environment; Healthcare Access and System Experience; and Food Environment. Each domain comprised three related indicators. Individual indicators were scored according to the degree of disadvantage (0 = no disadvantage, 1 = moderate disadvantage, and 2 = marked disadvantage), and the indicator scores within each domain were summed to generate a composite domain score. Higher scores represented a greater burden of adverse social determinants within that domain. Prespecified cut-offs were then applied to classify participants as having an adverse versus non-adverse SDOH status for each domain.*

### Scoring of SDOH variables

Indicator scoring and domain thresholds were specified a priori by the study team. DISDOH was subsequently used to support conceptual framing and interpretation rather than as the scoring instrument. Individual indicators were assigned ordinal burden scores of 0 (low burden), 1 (moderate burden), or 2 (high burden) with higher scores representing greater social disadvantage. Indicator scores were summed to generate domain-specific scores. In total, each of the six SDOH domains could add up to a possible score ranging from 0 to 6. For each domain, summed scores of 0–2 were classified as low burden, 3–4 as moderate burden, and 5–6 as high burden. The constituent indicators, together with an illustrative questionnaire item and their conceptual relationship to DISDOH, are summarized in table 1. The complete questions and detailed scoring rubric are presented in the Supplementary Appendix.

### Analysis of SDOH variables

For analysis, the six SDOH domain scores were used as the primary exposure variables. Each domain score was derived by summing the scores of its three constituent indicators and was analyzed as a domain-level measure of social burden.

### Psychological Health

Depressive symptoms, anxiety symptoms, and diabetes-related distress were assessed using the PHQ-9, GAD-7, and PAID scales, respectively [19–21]. The questionnaires were interviewer-administered by a trained data clerk in English or Kiswahili according to participant preference, with clarification provided when required. However, formal psychometric validation of the Kiswahili versions in this study population was not undertaken. The Patient Health Questionnaire-9 (PHQ-9), a nine-question tool, was used to ask about depressive symptoms. Each response was scored from 0 (not at all) to 3 (nearly every day), with total scores ranging from 0 to 27. Interpretation was as follows: 0–4 (none), 5–9 (mild), 10–14 (moderate), 15–19 (moderately severe), and 20–27 (severe depression) [20]. Participants reporting suicidal ideation on PHQ-9 item 9 were immediately referred to a psychologist for further assessment and management. The Generalized Anxiety Disorder-7 (GAD-7) instrument was used to assess anxiety symptoms. Scores ranged from 0 to 21 and categorized as: 0–4 (none), 5–9 (mild), 10– 14 (moderate), and 15–21 (severe anxiety) [20]. Scores of ≥10 on the PHQ-9 and GAD-7 were considered clinically significant. The Problem Areas in Diabetes (PAID) tool, a 20-item questionnaire explored questions relevant to distress in diabetes. Participants scored each item on a scale from 0 (not a problem) to 4 (a serious problem). Total scores were calculated and then multiplied by 1.25, yielding a score out of 100. A score of 40 or above was considered indicative of high diabetes distress. [21].

### Glycemic control, Albuminuria and Lipid profile

Glycemic control was measured using glycated hemoglobin (HbA1c) through high-performance liquid chromatography (HPLC). Glycemic control was categorized as at target (HbA1c <7.0%), above target (7.0–8.9%), and markedly above target (≥9.0%) [22]. Albuminuria was determined through a single test, urine albumin-to-creatinine ratio (UACR), using a random urine sample analyzed by Abbott Afinion 2 Analyzer through a colorimetric-based spectroscopy. Results were categorized as <30 mg/g (normal), 30–299 mg/g (moderately increased albuminuria: >300 mg/g (severely increased albuminuria) [23].

Lipid profile analysis comprised Low Density Lipoproteins (LDL) and High-Density Lipoproteins (HDL) Cholesterol biomarker. The LDL-Cholesterol was analyzed using the LDL-Cholesterol Gen. 3 (LDLC3) assay, an in vitro homogeneous enzymatic colorimetric test while the HDL-Cholesterol Gen. 4 (HDLC4) assay, an in vitro diagnostic test, directly measured HDL-cholesterol in serum. Results for LDL-C S were categorized as: Optimal: < 100 mg/dL (< 2.6 mmol/L); borderline high: 100–129 mg/dL (2.6–3.3 mmol/L). High (abnormal): ≥ 130 mg/dL (≥ 3.4 mmol/L). The HDL results were reported as low (abnormal): < 40 mg/dL (< 1.03 mmol/L); acceptable/Protective: ≥ 40 mg/dL (≥ 1.0 mmol/L); high (cardioprotective): ≥ 60 mg/dL (≥ 1.55 mmol/L) [24]. Additionally, LDL/HDL ratio was computed and categorized as low (<2.0), moderate (2.0–3.0), and high (>3.0) cardiovascular risk based on previously published cardiovascular risk stratification using lipoprotein ratios [24].

### Statistical analysis

Data were analysed using IBM SPSS Statistics version 28. Descriptive statistics were used to summarise sociodemographic, clinical, psychological, and social determinants of health characteristics. Categorical variables are presented as frequencies and percentages, while continuous variables are presented as means with standard deviations. Bivariate analyses included chi-square tests, independent-samples t-tests or one-way analysis of variance, and Pearson’s or Spearman’s correlation coefficients, according to variable type and distribution.

Multivariable regression analyses were used to examine associations between the six SDOH domain scores and glycaemic, renal, lipid, and psychological outcomes. HbA1c was categorised as <7.0%, 7.0–8.9%, and ≥9.0% and analysed as an ordered outcome using multivariable ordinal logistic regression. The resulting adjusted odds ratio therefore represents the change in the odds of being in a higher HbA1c category for each one-unit increase in the respective SDOH domain score.

Binary logistic regression was used for the remaining clinical and psychological outcomes. Elevated UACR was defined as ≥30 mg/g versus <30 mg/g; LDL cholesterol above target as ≥2.6 mmol/L versus <2.6 mmol/L; low HDL cholesterol as <1.03 mmol/L versus ≥1.03 mmol/L; and an elevated LDL/HDL ratio as ≥2.0 versus <2.0. Psychological outcomes were defined as clinically significant diabetes distress (PAID ≥40 versus <40), depressive symptoms (PHQ-9 ≥10 versus <10), and anxiety symptoms (GAD-7 ≥10 versus <10).

Separate multivariable models were fitted for each SDOH domain and outcome and adjusted for age, sex, and duration of diabetes. Associations are reported as adjusted odds ratios (aORs) with 95% confidence intervals. Statistical significance was defined as a two-sided p-value <0.05.

## Results

Table 2 summarizes the characteristics of the 164 participants. The mean age was 24.96 ± 4.35 years, with 99 (60.4%) participants aged 18–25 years, and the mean diabetes duration was 7.26 ± 5.70 years. Despite 108 (65.9%) participants receiving basal–bolus insulin therapy, glycaemic control was markedly poor. Participants had a mean HbA1c of 11.50 ± 3.44% with 134 (81.7%) having HbA1c ≥9.0%, while only 8 (4.9%) achieved HbA1c <7.0%. Psychological morbidity was substantial, with diabetes distress, clinically significant depressive symptoms, and clinically significant anxiety present in 48 (29.3%), 45 (27.5%), and 29 (17.6%) participants, respectively. Early cardio-renal complication markers were also frequent: 46 (28.0%) participants had a single elevated UACR, while 92 (56.1%) participants had LDL-C above 2.6 mmol/L and 105 (64.0%) participants had an LDL/HDL ratio ≥2.

**Table 2.**
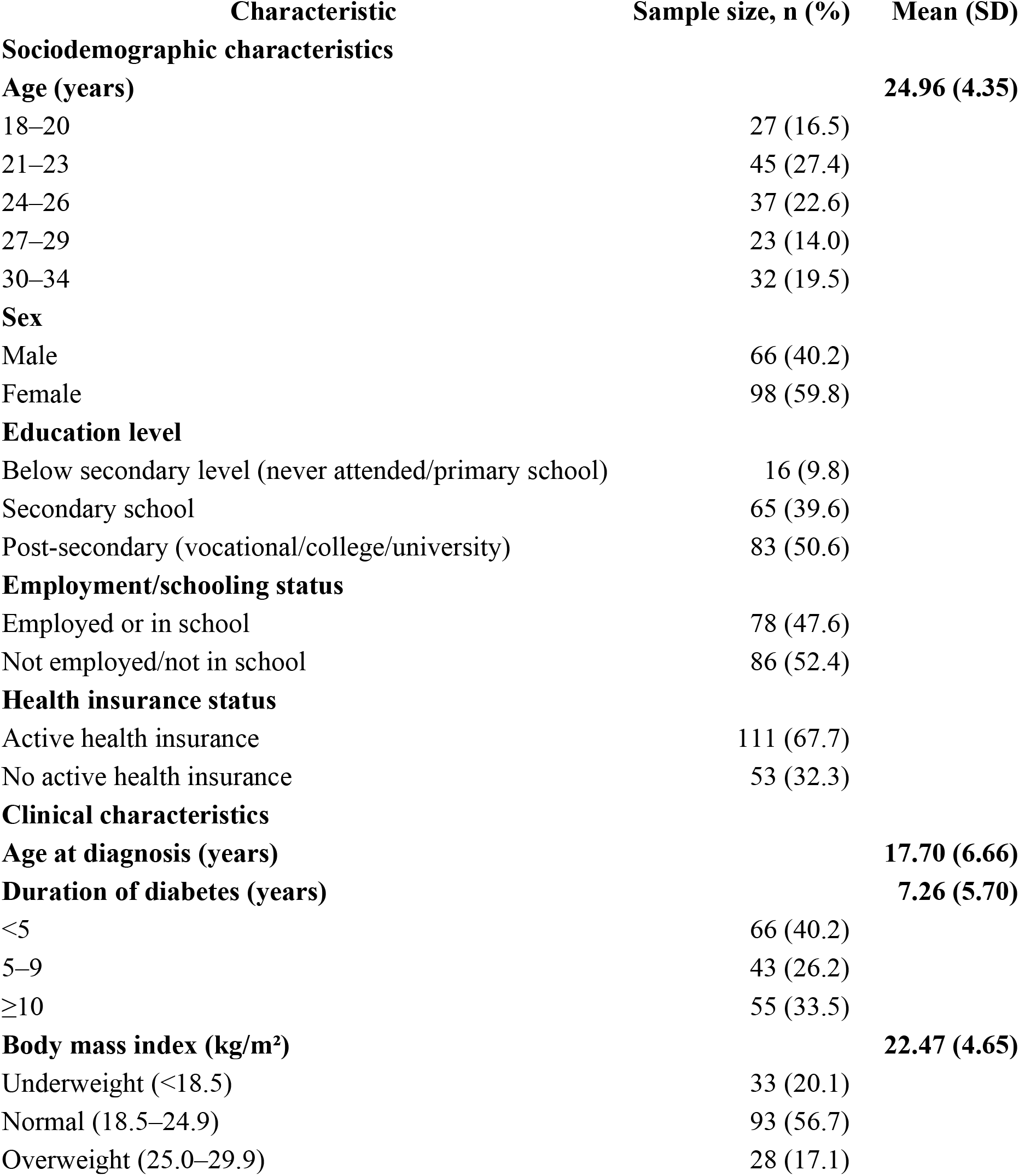
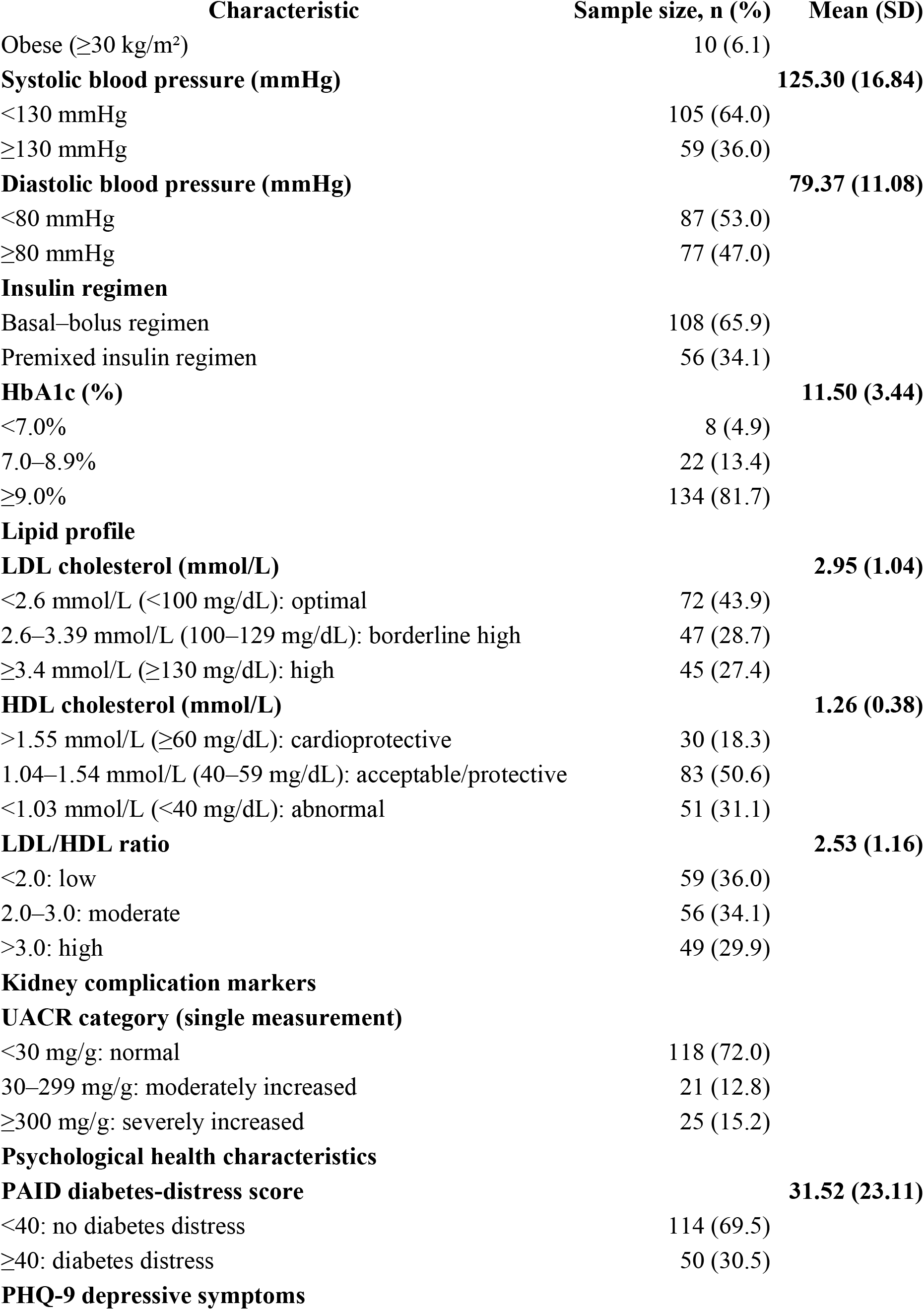
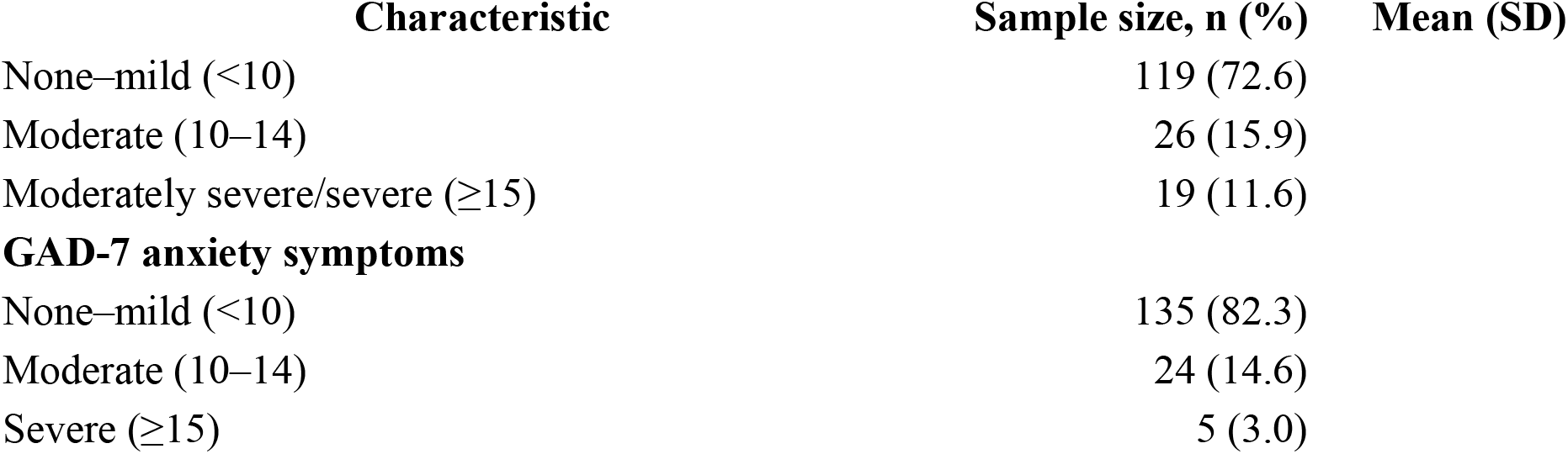
Baseline Sociodemographic, Clinical, Psychological, and Social Determinants of Health Characteristics of Young Adults Living with Type 1 Diabetes in Kenya (N = 164)

| **Characteristic** | **Sample size, n (%)** | **Mean (SD)** |
|---|---|---|
| **Sociodemographic characteristics** |  |  |
| **Age (years)** |  | **24.96 (4.35)** |
| 18–20 | 27 (16.5) |  |
| 21–23 | 45 (27.4) |  |
| 24–26 | 37 (22.6) |  |
| 27–29 | 23 (14.0) |  |
| 30–34 | 32 (19.5) |  |
| **Sex** |  |  |
| Male | 66 (40.2) |  |
| Female | 98 (59.8) |  |
| **Education level** |  |  |
| Below secondary level (never attended/primary school) | 16 (9.8) |  |
| Secondary school | 65 (39.6) |  |
| Post-secondary (vocational/college/university) | 83 (50.6) |  |
| **Employment/schooling status** |  |  |
| Employed or in school | 78 (47.6) |  |
| Not employed/not in school | 86 (52.4) |  |
| **Health insurance status** |  |  |
| Active health insurance | 111 (67.7) |  |
| No active health insurance | 53 (32.3) |  |
| **Clinical characteristics** |  |  |
| **Age at diagnosis (years)** |  | **17.70 (6.66)** |
| **Duration of diabetes (years)** |  | **7.26 (5.70)** |
| <5 | 66 (40.2) |  |
| 5–9 | 43 (26.2) |  |
| ≥10 | 55 (33.5) |  |
| **Body mass index (kg/m<sup>2</sup>)** |  | **22.47 (4.65)** |
| Underweight (<18.5) | 33 (20.1) |  |
| Normal (18.5–24.9) | 93 (56.7) |  |
| Overweight (25.0–29.9) | 28 (17.1) |  |
| **Characteristic** | **Sample size, n (%)** | **Mean (SD)** |
| Obese (≥30 kg/m<sup>2</sup>) | 10 (6.1) |  |
| **Systolic blood pressure (mmHg)** |  | **125.30 (16.84)** |
| <130 mmHg | 105 (64.0) |  |
| ≥130 mmHg | 59 (36.0) |  |
| **Diastolic blood pressure (mmHg)** |  | **79.37 (11.08)** |
| <80 mmHg | 87 (53.0) |  |
| ≥80 mmHg | 77 (47.0) |  |
| **Insulin regimen** |  |  |
| Basal–bolus regimen | 108 (65.9) |  |
| Premixed insulin regimen | 56 (34.1) |  |
| **HbA1c (%)** |  | **11.50 (3.44)** |
| <7.0% | 8 (4.9) |  |
| 7.0–8.9% | 22 (13.4) |  |
| ≥9.0% | 134 (81.7) |  |
| **Lipid profile** |  |  |
| **LDL cholesterol (mmol/L)** |  | **2.95 (1.04)** |
| <2.6 mmol/L (<100 mg/dL): optimal | 72 (43.9) |  |
| 2.6–3.39 mmol/L (100–129 mg/dL): borderline high | 47 (28.7) |  |
| ≥3.4 mmol/L (≥130 mg/dL): high | 45 (27.4) |  |
| **HDL cholesterol (mmol/L)** |  | **1.26 (0.38)** |
| >1.55 mmol/L (≥60 mg/dL): cardioprotective | 30 (18.3) |  |
| 1.04–1.54 mmol/L (40–59 mg/dL): acceptable/protective | 83 (50.6) |  |
| <1.03 mmol/L (<40 mg/dL): abnormal | 51 (31.1) |  |
| **LDL/HDL ratio** |  | **2.53 (1.16)** |
| <2.0: low | 59 (36.0) |  |
| 2.0–3.0: moderate | 56 (34.1) |  |
| >3.0: high | 49 (29.9) |  |
| **Kidney complication markers** |  |  |
| **UACR category (single measurement)** |  |  |
| <30 mg/g: normal | 118 (72.0) |  |
| 30–299 mg/g: moderately increased | 21 (12.8) |  |
| ≥300 mg/g: severely increased | 25 (15.2) |  |
| **Psychological health characteristics** |  |  |
| **PAID diabetes-distress score** |  | **31.52 (23.11)** |
| <40: no diabetes distress | 114 (69.5) |  |
| ≥40: diabetes distress | 50 (30.5) |  |
| **PHQ-9 depressive symptoms** |  |  |
| **Characteristic** | **Sample size, n (%)** | **Mean (SD)** |
| None–mild (<10) | 119 (72.6) |  |
| Moderate (10–14) | 26 (15.9) |  |
| Moderately severe/severe (≥15) | 19 (11.6) |  |
| **GAD-7 anxiety symptoms** |  |  |
| None–mild (<10) | 135 (82.3) |  |
| Moderate (10–14) | 24 (14.6) |  |
| Severe (≥15) | 5 (3.0) |  |

The SDOH are presented in the heat map figure 2 below.

**Figure 2:**
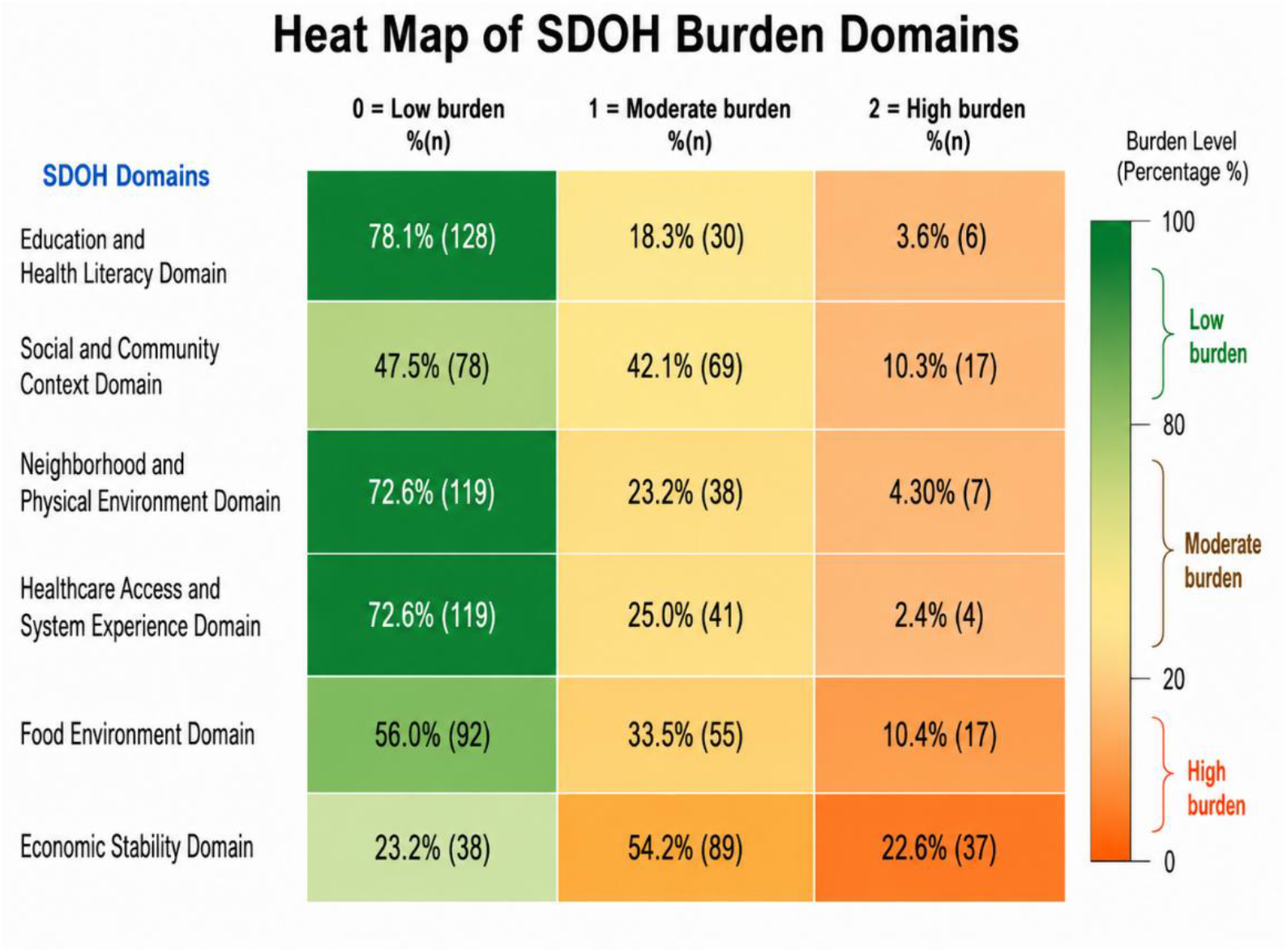
Distribution of social determinants of health (SDOH) burden across six domains. Each SDOH domain is comprised of three indicators. For example, the “healthcare access and system experience” domain includes access to multidisciplinary care, health insurance, and clinic experience as described in the methodology.

Figure 2 shows marked variation in SDOH burden across the six domains. Economic stability represented the greatest social vulnerability, with 89 (54.2%) participants experiencing moderate burden and 37 (22.6%) high burden, corresponding to 76.8% with moderate-to-high burden overall. Moderate-to-high burden was also common in the social and community context [86 (52.4%)] and food environment [72 (43.9%)] domains. In contrast, most participants had low burden in education and health literacy [128 (78.1%)], neighbourhood and physical environment [119 (72.6%)], and healthcare system [119 (72.6%)] domains. High burden was most frequent for economic stability [37 (22.6%)], compared with food environment [17 (10.4%)] and social and community context [17 (10.3%)], highlighting economic vulnerability as the predominant adverse social determinant in this cohort.

Table 3. Multivariable associations of age, duration of diabetes, and HbA1c category with renal, lipid, and psychological outcomes. Values are presented as adjusted odds ratios (aORs) with 95% confidence intervals (CIs), with corresponding p-values shown on the second line. Bold indicates statistical significance at p<0.05. Age and duration of diabetes were analysed as continuous variables, and HbA1c as an ordinal variable with three categories, <7.0%, 7.0–8.9%, and ≥9.0%. The corresponding aOR therefore represents the odds associated with moving one HbA1c category higher, or more. Psychological outcomes were defined as diabetes distress (PAID >40), clinically significant depressive symptoms (PHQ-9 ≥10), and clinically significant anxiety symptoms (GAD-7 ≥10). Statistical significance was defined as p<0.05.

**Table 3.** Multivariable associations of age, duration of diabetes, and HbA1c with clinical and psychosocial outcomes.

| **Independent variable** | **Elevated UACR** | **LDL above target** | **Low HDL** | **Elevated LDL/HDL ratio** | **Diabetes distress (PAID ≥40)** | **Depressive symptoms (PHQ-9 ≥10)** | **Anxiety symptoms (GAD-7 ≥10)** |
|---|---|---|---|---|---|---|---|
| **Age** | 1.02 (0.94–1.11) *p*=0.597 | 0.94 (0.87–1.01) *p*=0.111 | 1.02 (0.94–1.11) *p*=0.575 | 0.97 (0.90–1.04) *p*=0.392 | 1.08 (1.00–1.17) *p*=0.052 | 1.03 (0.92–1.14) *p*=0.629 | 1.04 (0.94–1.16) *p*=0.447 |
| **Duration of diabetes** | **1.10 (1.03–1.17) *p*=0.003** | 1.06 (1.00–1.12) *p*=0.052 | **0.91 (0.85–0.97) *p*=0.007** | 0.98 (0.93–1.04) *p*=0.571 | 0.96 (0.90–1.02) *p*=0.183 | 1.02 (0.94–1.10) *p*=0.657 | 1.00 (0.93–1.09) *p*=0.932 |
| **HbA1c category** | 1.48 (0.69–3.18) *p*=0.314 | 1.48 (0.79–2.77) *p*=0.223 | 1.07 (0.55–2.08) *p*=0.847 | 1.71 (0.93–3.15) *p*=0.084 | 1.17 (0.60–2.28) *p*=0.641 | 0.84 (0.37–1.92) *p*=0.686 | 1.34 (0.51–3.51) *p*=0.552 |

In multivariable logistic regression, longer duration of diabetes was significantly associated with elevated UACR, with each additional year of diabetes corresponding to approximately 10% higher adjusted odds of elevated UACR (aOR 1.10, 95% CI 1.03–1.17; *p*=0.003). Longer diabetes duration was also associated with lower odds of low HDL cholesterol (aOR 0.91, 95% CI 0.85–0.97; *p*=0.007), while its association with LDL cholesterol above target was borderline (aOR 1.06, 95% CI 1.00–1.12; *p*=0.052). No significant associations were observed between age and the renal, lipid, or psychological outcomes, although age showed a borderline association with diabetes distress (aOR 1.08, 95% CI 1.00–1.17; *p*=0.052). Increasing HbA1c category was not significantly associated with elevated UACR, LDL, low HDL, diabetes distress, depressive symptoms, or anxiety symptoms, although there was a non-significant trend toward higher odds of an elevated LDL/HDL ratio with worsening HbA1c category (aOR 1.71, 95% CI 0.93–3.15; *p*=0.084).

In adjusted analyses, in figure 3, none of the six SDOH domains showed a statistically significant association with the HbA1c outcome. Nevertheless, all aORs were greater than 1.0, suggesting a consistent direction toward higher odds of an adverse HbA1c outcome with increasing social disadvantage. The largest associations were observed for education and health literacy (aOR 1.36, 95% CI 0.83–2.25; *p*=0.223), social and community context (aOR 1.29, 95% CI 0.84–1.99; *p*=0.254), and healthcare access and system experience (aOR 1.27, 95% CI 0.88–1.84; *p*=0.209). Smaller non-significant associations were observed for food environment and household food security (aOR 1.17, 95% CI 0.83–1.65; *p*=0.375), economic stability (aOR 1.13, 95% CI 0.76–1.68; *p*=0.551), and neighbourhood and physical environment (aOR 1.04, 95% CI 0.71–1.52; *p*=0.833).

**Figure 3.**
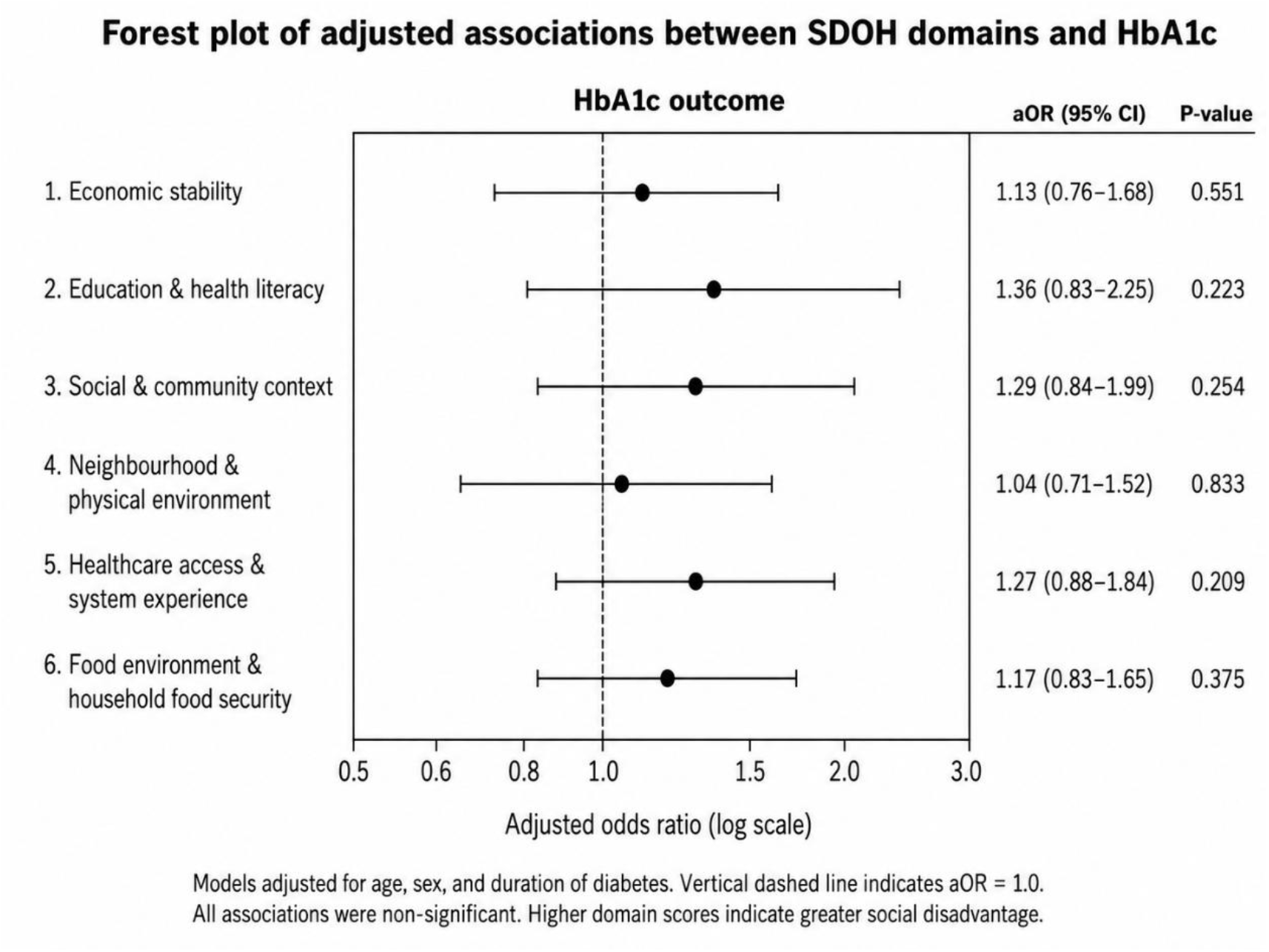
Adjusted associations between social determinants of health domains and the primary outcome: HbA1c. Forest plot showing adjusted odds ratios (aORs) and 95% confidence intervals for associations between the six SDOH domains and the HbA1c outcome. Models were adjusted for age, sex, and duration of diabetes. Higher SDOH domain scores indicate greater social disadvantage. The vertical dashed line represents the null value (aOR = 1.0).

Greater economic instability was associated with 40% higher adjusted odds of elevated UACR (aOR 1.40, 95% CI 1.05–1.85; *p*=0.020) as shown in figure 4. The other SDOH domains showed a positive association but were not significantly associated with elevated UACR, including education and health literacy (aOR 1.08, 95% CI 0.77–1.53; *p*=0.647), social and community context (aOR 1.24, 95% CI 0.92–1.67; *p*=0.160), neighbourhood and physical environment (aOR 1.04, 95% CI 0.80–1.35; *p*=0.779), healthcare access and system experience (aOR 1.11, 95% CI 0.86–1.43; *p*=0.415), and food environment and household food security (aOR 1.12, 95% CI 0.88–1.42; *p*=0.355).

**Figure 4.**
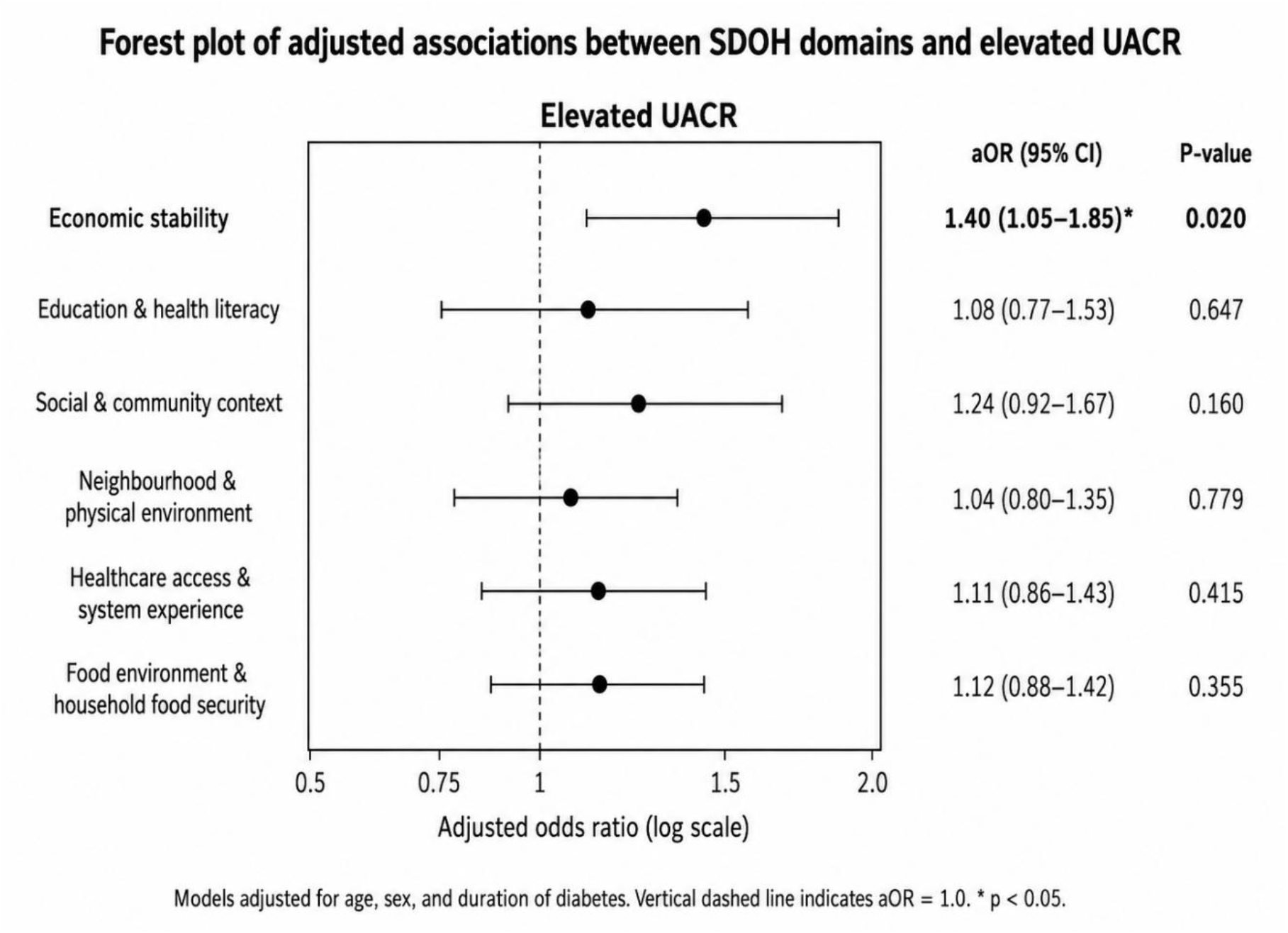
Forest plot of adjusted associations between social determinants of health domains and the secondary outcome: elevated UACR. Forest plot showing adjusted odds ratios (aORs) and 95% confidence intervals for the associations between the six SDOH domains and elevated UACR. Models were adjusted for age, sex, and duration of diabetes. Higher SDOH domain scores indicate greater social disadvantage. The vertical dashed line represents the null value (aOR = 1.0).

In multivariable logistic regression adjusted for age, sex, and duration of diabetes, in figure 5, no SDOH domain was significantly associated with the LDL cholesterol outcome (all *p*>0.05). Similarly, no significant associations were observed for HDL cholesterol, although greater food environment and household food security burden showed a borderline association (aOR 1.25, 95% CI 0.99–1.57; *p*=0.059). For the LDL/HDL ratio, greater neighbourhood and physical environment burden was significantly associated with higher odds of an elevated LDL/HDL ratio (aOR 1.33, 95% CI 1.03–1.73; *p*=0.030). Economic stability (aOR 1.14, 95% CI 0.94–1.36; *p*=0.210), education and health literacy (aOR 1.07, 95% CI 0.77–1.47; *p*=0.695), social and community context (aOR 0.93, 95% CI 0.71–1.22; *p*=0.608), healthcare access and system experience (aOR 0.88, 95% CI 0.70–1.12; *p*=0.304), and food environment and household food security (aOR 1.14, 95% CI 0.92–1.42; *p*=0.230) were not significantly associated with the LDL/HDL ratio.

**Figure 5.**
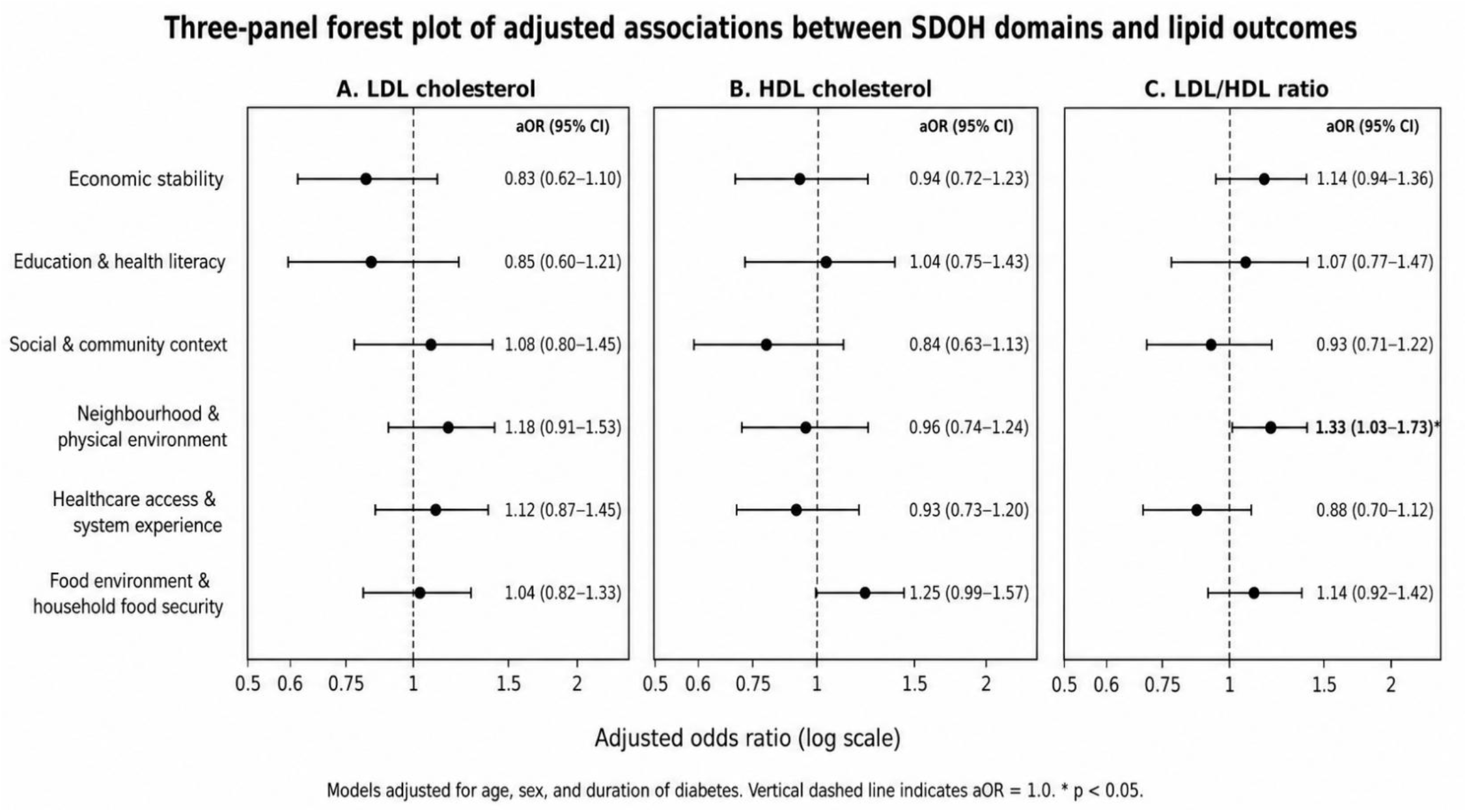
Multivariable logistic regression analysis of associations between social determinants of health domains and the secondary outcome: dyslipidemia. Forest plots show adjusted odds ratios (aORs) and 95% confidence intervals for associations between the six SDOH domains and (A) LDL cholesterol, (B) HDL cholesterol, and (C) LDL/HDL ratio. Models were adjusted for age, sex, and duration of diabetes. Higher SDOH domain scores indicate greater social disadvantage. The vertical dashed line represents the null value (aOR = 1.0).

Greater SDOH burden was associated with several adverse psychological outcomes in figure 6. For diabetes distress, higher burden in economic stability (aOR 1.42, 95% CI 1.17–1.72), neighbourhood and physical environment (aOR 1.39, 95% CI 1.07–1.80), healthcare access and system experience (aOR 1.45, 95% CI 1.12–1.87), and food environment and household food security (aOR 1.30, 95% CI 1.03–1.64) was significantly associated with higher odds of distress. No SDOH domain was significantly associated with depressive symptoms, although social and community context and food-environment burden showed non-significant trends toward higher odds. Anxiety symptoms were significantly associated with greater social and community context burden (aOR 1.70, 95% CI 1.14–2.53), healthcare access and system experience burden (aOR 1.68, 95% CI 1.19–2.36), and economic instability (aOR 1.31, 95% CI 1.02–1.68).

**Figure 6.**
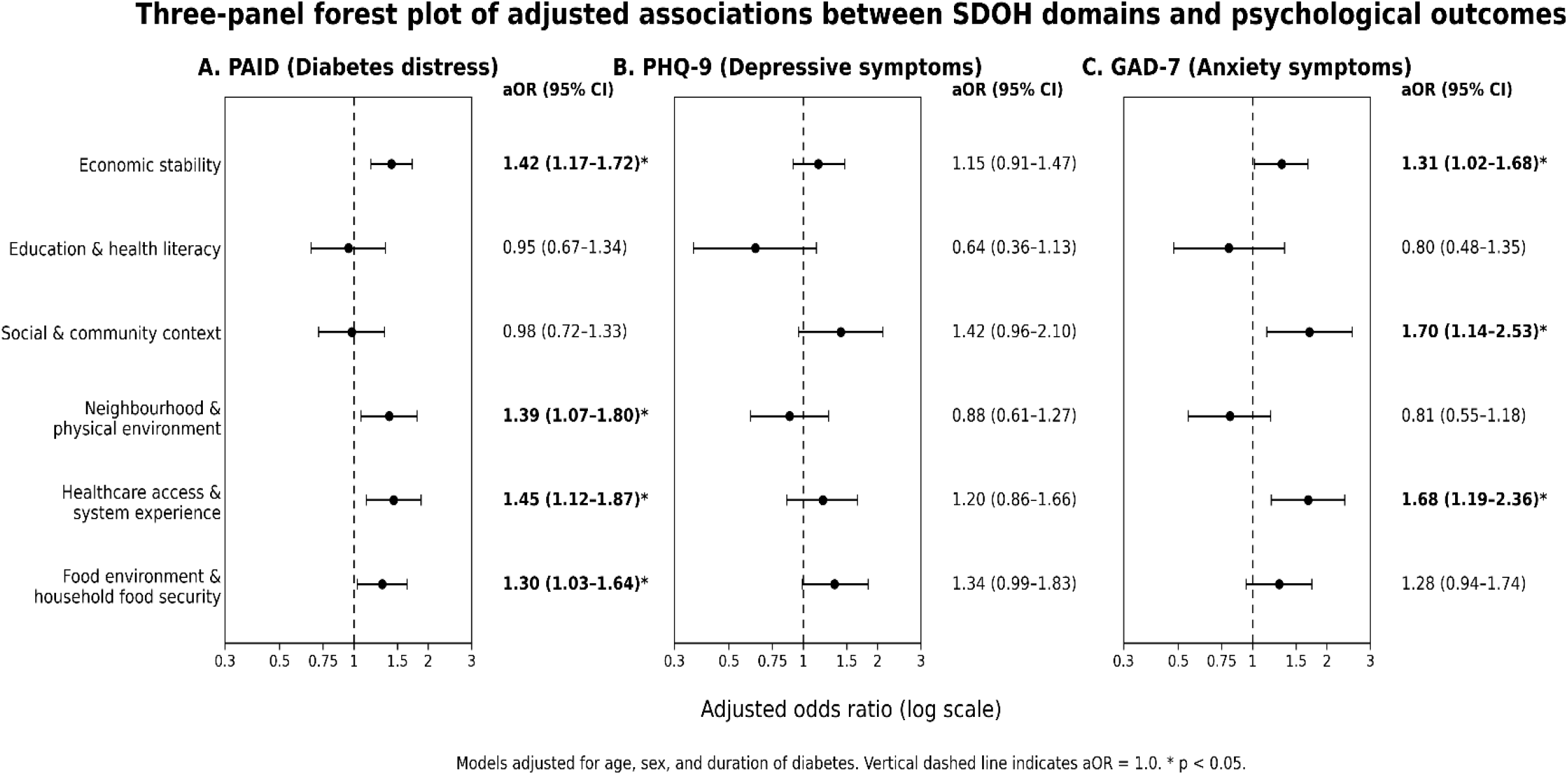
Adjusted associations between social determinants of health domains and the secondary outcome: psychological health. Three-panel Forest plots show adjusted odds ratios (aORs) and 95% confidence intervals for associations between the six social determinants of health (SDOH) domains and (A) diabetes distress assessed using PAID, (B) depressive symptoms assessed using PHQ-9, and (C) anxiety symptoms assessed using GAD-7. Separate multivariable logistic regression models were fitted for each SDOH domain and adjusted for age, sex, and duration of diabetes. Higher SDOH domain scores indicate greater social disadvantage. The vertical dashed line represents the null value (aOR = 1.0).

## Discussion

This study demonstrates the interconnected burden of social disadvantage, psychological morbidity, poor glycaemic control, and early complication risk among young adults with type 1 diabetes in Kenya. Although higher SDOH burden generally showed a positive but non-significant trend with poor glycaemic control, social and structural barriers were more clearly related to diabetes distress and anxiety, suggesting that their effects may operate partly through psychological and healthcare pathways rather than directly through HbA1c alone. Economic instability was associated with higher odds of albuminuria while neighbourhood and physical-environment disadvantage positively correlated with an adverse LDL/HDL ratio. These findings support a broader model of T1D care that addresses social vulnerability and mental health alongside conventional metabolic management.

A substantial SDOH burden was evident across all six domains, underscoring the multiple social and structural pressures faced by young adults living with T1D. The greatest SDOH burden was seen in economic stability, where 76.8% of participants had moderate-to-high burden followed by social and community context at 52.4%. This suggests that limited social support, financial insecurity, unemployment or unstable work, and dependence on others, individual components of the economic stability domain, may be particularly important challenges during early adulthood. Food environment also showed a notable moderate burden (43.9%), reflecting the potential difficulty of matching insulin with carbohydrates and availability of healthy snacks to prevent hypoglycemia [25–27]. Although education and health literacy and neighbourhood and physical environment showed comparatively lower overall burden, these domains still represent important barriers for a subset of participants. These findings are consistent with studies among underserved populations globally, which have reported challenges related to socioeconomic status, food insecurity, education, and healthcare access [25–28]. The observed burden support integrated action by clinicians, policymakers, and community advocates to routinely identify and address financial, social, and food-related barriers as part of comprehensive T1D care for young adults.

Glycaemic outcomes were particularly concerning, with more than 90% of participants having HbA1c levels above recommended targets and a mean HbA1c of 11.5%. These findings are consistent with studies from sub-Saharan Africa reporting poor glycaemic control, with HbA1c levels often exceeding 10% [29–31]. However, glycaemic control in our cohort was poorer than global estimates among children, adolescents and youth, where mean HbA1c levels were 8.23%, 8.73%, 9.20%, and 10.11% in high-income, upper-middle-income, lower-middle-income, and low-income countries, respectively [32, 33]. Poorer glycaemic control among participants aged 18–25 years is also consistent with evidence that adolescents and young adults often experience worse glycaemic outcomes than other age groups.

All six adverse SDOH domains showed a non-significant trend toward higher HbA1c, consistent with the variable associations reported in previous diabetes studies [33]. This suggests that social vulnerabilities may not independently determine glycaemic control but may influence it indirectly through other ways like treatment adherence [29], self-management and caregiver support [34], healthcare engagement, psychological well-being [35], and diabetes literacy. The uniformly poor glycaemic control in this cohort may also have reduced the ability to distinguish differences across SDOH burden levels. These findings support addressing adverse social determinants alongside intensified diabetes management, as improving glycaemic control in young adults with T1D may require not only optimizing insulin therapy but also reducing financial, social, and food-related barriers to self-management.

Psychological burden was considerable, with nearly 30% of participants reporting clinically significant diabetes distress and depressive symptoms. The prevalence of diabetes distress in our study was substantially higher than that reported in one high-income study setting by Albright et al., who found diabetes distress and depression in 3.6% and 5% of youth with T1D, respectively [36]. However, direct comparison should be cautious given differences in population and care-related factors, as participants in Albright et al. were younger (mean age 17.3 years), had lower mean HbA1c (8.6%), and most used insulin pumps and CGM. Our findings were more comparable to those reported among adolescents and young people with T1D in other low, middle and certain high-income settings, where a substantial psychological burden has also been described [37]. Adverse SDOH domains were more consistently associated with psychological outcomes than with HbA1c. Economic instability, food environment burden, healthcare barriers, and neighbourhood and physical environment burden were associated with higher diabetes distress, while anxiety was associated with greater social and community context and healthcare system burden. Adverse SDOH may increase the complexity of diabetes self-management by limiting access to insulin, glucose monitoring, transport, nutritious food,healthcare support, and social support while increasing financial stress and uncertainty. These cumulative demands may contribute to diabetes distress and anxiety even when clinical care is available [38–40]. The stronger associations observed between SDOH and psychological well-being than with HbA1c suggest that adverse social conditions may influence diabetes outcomes through psychological and behavioural pathways before manifesting as measurable differences in glycaemic control. Our results support integrating routine screening for diabetes distress, depression, and anxiety into T1D care, alongside psychosocial support and interventions addressing underlying social and structural barriers.

Approximately 28% of participants had elevated UACR, suggesting a substantial burden of possible early diabetic kidney involvement. This proportion was comparable to a Botswana study among individuals younger than 24 years, which reported albuminuria in 28.3% [41], but higher than findings from Tanzania [42], Alleyn et al. (23%) [42], and a systematic review by Stefano et al., which reported a global mean albuminuria prevalence of 9.5% among children and adolescents [46]. This difference may partly reflect the older age, longer diabetes duration, and poorer glycaemic control of our cohort. Elevated UACR was strongly associated with longer diabetes duration, underscoring prolonged disease exposure as an important contributor to early kidney injury [45]. Chronic hyperglycaemia contributes to glomerular injury through haemodynamic changes, oxidative stress, inflammation, and renal microvascular damage, thereby increasing the risk of albuminuria and nephropathy [4]. Economic instability was also significantly associated with elevated UACR, suggesting that socioeconomic vulnerability may contribute to renal risk through reduced access to care, delayed detection, and cumulative metabolic and psychosocial burden [46]. However, this association should be interpreted cautiously because UACR was measured only once and the cross-sectional design does not establish temporality. The high burden of possible early kidney involvement supports routine UACR screening in young adults with T1D, alongside improved glycaemic management and strategies to reduce socioeconomic barriers that may delay detection and access to renal-protective care.

Dyslipidaemia was also common, with more than half of participants having LDL-C above 2.6 mmol/L. This burden was higher than that reported in other studies, including the SEARCH study among youth with T1D [47,48], and may partly reflect the poor glycaemic control observed in this cohort. Longer diabetes duration showed a borderline association with LDL above target (aOR 1.06, p=0.052), while higher HbA1c showed a non-significant trend toward an elevated LDL/HDL ratio (aOR 1.71, p=0.084). Conversely, longer diabetes duration was associated with lower odds of low HDL (aOR 0.91, p=0.007). Although seemingly counterintuitive, higher HDL-C with increasing diabetes duration has been reported in some cohorts of young people with diabetes [44]. HDL-C levels in T1D may remain relatively preserved or elevated, potentially reflecting effects of exogenous insulin on lipoprotein metabolism [49, 50]. However, evidence is inconsistent, and the clinical significance of this finding requires further investigation. Overall, these findings suggest that young adults in this setting are accumulating cardiovascular risk factors early in adulthood. The association between neighbourhood and physical environment burden and elevated LDL/HDL ratio further suggests that adverse social and environmental conditions may contribute to early cardiovascular risk, although confirmation in longitudinal studies is needed. The early accumulation of dyslipidaemia and its association with social and environmental burden supports routine lipid screening and cardiovascular risk reduction in young adults with T1D, alongside interventions that address adverse SDOH and improve glycaemic control.

This study highlights major unmet needs among young adults with T1D in Kenya and suggests that insulin access alone is insufficient. Integrated care should address biomedical, psychosocial, and social needs, with routine assessment of SDOH and mental health alongside glycaemic monitoring, particularly during transition to adult care. Young adults with high financial, food, healthcare, or social-support burden may benefit from targeted interventions such as strengthened peer support, mental health services, multidisciplinary care, financial protection, and improved access to essential self-management resources. The high prevalence of poor glycaemic control, psychological distress, dyslipidaemia, and single elevated UACR also supports earlier surveillance for renal and cardiovascular risk. These findings suggest that improving insulin access alone may be insufficient to achieve optimal long-term outcomes without simultaneously addressing broader social and psychological barriers.

Unlike previous studies that have examined glycaemic control, psychological health or individual social determinants in isolation, this study simultaneously evaluated multiple domains of social disadvantage, psychological well-being, glycaemic control, kidney risk, and dyslipidaemia among young adults with T1D in a sub-Saharan African setting. This integrated approach provides a more comprehensive understanding of how social, psychological, and biological factors interact to influence diabetes outcomes in young adults with T1D. Importantly, this study is among the first to apply a comprehensive framework to assess the relationship between multiple non-clinical determinants and T1D outcomes in this population. Future longitudinal and implementation studies should evaluate whether interventions targeting modifiable social determinants improve psychological well-being, metabolic outcomes, and long-term complications among young adults living with T1D in resource-limited settings. Such studies should also confirm persistent albuminuria through repeat testing and evaluate context-specific SDOH instruments prospectively.

## Limitations

This study has several limitations. Its cross-sectional design precludes causal inference. Non-probability recruitment may have introduced selection bias and limited generalizability. Nevertheless, it provides important baseline evidence in a data-scarce population of young adults with T1D. Diabetes type was determined clinically because routine islet autoantibody and C-peptide testing is not widely available in our resource limited setting, creating some risk of misclassification, particularly among those diagnosed in adulthood. The Kiswahili versions of PAID, PHQ-9, and GAD-7 were not formally validated in this population, and interviewer-administered and self-reported variables may be subject to recall or social desirability bias. The context-specific SDOH indices were based on established frameworks and local evidence but require formal validation. Although the overall sample size was adequately powered for the primary analyses, some subgroup analyses, particularly among participants with optimal HbA1c, had small numbers and may have lacked power. UACR was measured once because resource constraints did not allow repeat testing; while this may overestimate possible early kidney involvement, a single elevated UACR remains clinically meaningful and provides important early renal risk data in a setting where such evidence is limited. Despite these limitations, the study provides a comprehensive assessment of social, psychological, glycaemic, renal, and cardiovascular risk factors in an understudied population of young adults with T1D. It also provides key incipient evidence in this setting that currently primarily centers insulin as a cornerstone for care.

## Statements and Declarations Ethical considerations

Ethical approval was obtained from the Kenyatta National Hospital–University of Nairobi Ethics and Research Committee (P595/06/2025). All study procedures were conducted in accordance with applicable ethical standards and regulations.

## Consent to participate

Written informed consent was obtained from all participants before enrolment in the study.

## Consent for publication

Not applicable. The manuscript does not contain identifiable individual participant information, images, or videos.

## Data Availability

All data produced in the present study are available upon reasonable request to the authors

## Data availability statement

The data supporting the findings of this study are available from the corresponding author upon reasonable request, subject to ethical and institutional requirements governing participant confidentiality and data sharing.

## Author Contributions

**Phoebe Wamalwa:** Conceptualization; Methodology; Investigation; Data curation; Formal analysis; Writing original draft; review and editing; Project administration.

**Cyrus Mugo:** Conceptualization; Methodology; Formal analysis; Supervision; review and editing of the manuscript.

**Christine Ngaruiya:** Conceptualization; Methodology; Supervision; Formal analysis, review and editing of manuscript.

All authors reviewed and approved the final manuscript.

